# Impact of the HPV vaccination programme on anal HPV infection in gay, bisexual and other men who have sex with men: multi-site enhanced surveillance study in specialist sexual health services in England

**DOI:** 10.64898/2026.06.29.26354430

**Authors:** Marta Checchi, Kavita Panwar, Clare E French, Hamish Mohammed, Simon Beddows, Matthew Hickman, Kate Soldan

## Abstract

**Background:** A national human papillomavirus (HPV) vaccination programme for gay, bisexual, and other men who have sex with men (GBMSM) in England began in 2016. We evaluated the impact of this programme by monitoring the prevalence of type-specific HPV infections over time.

**Methods:** Residual rectal swab specimens were collected from GBMSM aged ≤45 years undergoing chlamydia and/or gonorrhoea screening at 10 SHS between 2017-19 (Phase 1) and 2021-24 (Phase 2). Specimens were linked to data on sociodemographic characteristics, previous STI diagnoses, and HPV vaccination reported to UKHSA’s national STI surveillance system. Anonymised specimens were tested for type-specific HPV DNA using an in-house multiplex PCR and Luminex-based genotyping test. We compared the prevalence of vaccine-type HPV in each phase, and in those with and without any reported HPV vaccinations.

**Results:** Data from 5,787 rectal swab specimens with HPV testing results were analysed: 3,291 in Phase 1 and 2,496 in Phase 2. There was a 36% decline in the prevalence of anal HPV16/18 infection between Phase 1 and Phase 2. Overall, the prevalence of HPV16/18 was similar in GBMSM with (16.6%; 95% CI 15.2-18.1) and without reported HPV vaccination (15.8%; 95% CI 14.5-17.1).

**Conclusion:** Eight years after HPV vaccination of GBMSM began, the prevalence of vaccine-type HPV in GBMSM has declined. Vaccine effectiveness estimates were uncertain, likely due to the combined effects of underreporting of vaccinations, herd protection, and bias in vaccination uptake. These findings, albeit ecological, bode well for this targeted programme meeting its aim of reducing HPV-related diseases amongst GBMSM and reducing health inequalities.

## INTRODUCTION

High-risk human papillomavirus (HPV) type infections can cause cancers of the cervix, anus, penis, vulva, vagina, and oropharynx [1]. HPV16 and HPV18 are detected in 80-90% of anal cancer cases worldwide, as in cervical cancer [2]. HPV types 6 and 11 cause approximately 90% of anogenital warts (AGW) in both males and females [3]. Gay, bisexual, and other men who have sex with men (GBMSM), particularly those who are living with HIV, have a higher risk of HPV acquisition and have been disproportionately affected by HPV-associated disease [4]. A systematic review estimated global pooled HPV prevalence among GBMSM of 78.4% at the anal site, 36.2% penile, 17.3% oral, and 15.4% urethral [5]. Despite lower incidence rates of anal cancers in men than in women overall (1.8 and 3.1 per 100,000 in males and females, respectively in England 2017-2019) [6], the incidence of anal cancer among GBMSM, particularly GBMSM living with HIV, is disproportionately high compared to heterosexual men (and women) and has been increasing annually [4]. A meta-analysis found anal cancer incidence rates of 19.0, 85.0, and 32.0 per 100,000 person-years in HIV-negative GBMSM, GBMSM living with HIV, and heterosexual men living with HIV, respectively [7].

A national HPV vaccination programme was introduced in the United Kingdom (UK) for females in September 2008 and extended to include males in 2019. The programme offers routine HPV vaccination to year 8 adolescents (aged 12–13 years) in schools, with catch-up eligibility up to age 25. Since the introduction of this programme, there has been evidence of substantial declines in the prevalence of vaccine type HPV infections in vaccine-eligible sexually active young females, as well as evidence of declines in AGW in this age group [8, 9].

There is good evidence that high coverage amongst females, as in the UK, confers indirect, herd protection to heterosexual males. However, GBMSM are expected to benefit far less from female vaccination [10]. Additionally, the risk of infection (and related disease) and the age-distribution of infection are different for GBMSM [11]. A randomised controlled trial found a vaccine efficacy of 95.8% against HPV16/18 persistent anal infection and HPV DNA detection in GBMSM in the per-protocol population (seronegative and HPV DNA negative swab at enrolment) [12]. Mathematical modelling suggested favourable cost-effectiveness for selective quadrivalent vaccination of GBMSM aged ≤40 years attending specialist SHSs and HIV clinics [10]. Based on this evidence, and to address health inequality within HPV control, the Joint Committee on Vaccination and Immunisation (JCVI) recommended a targeted HPV vaccination programme for GBMSM aged ≤45 years attending specialist SHSs and HIV clinics, subject to cost-effective vaccine procurement and delivery of the programme and minimal disruption to clinics. Implementation started with a pilot programme which ran from June 2016 to April 2018 in 42 specialist SHSs and HIV clinics in England [13]. National roll-out followed, with all clinics implementing vaccination as of April 2019. The programme started with a 3-dose course for all and is now (since 2023) 1-dose for individuals aged under 25 years, 2-dose for individuals aged 25 to 45 years (inclusive) and 3-doses for individuals known to be immunocompromised and/or living with HIV at the time of vaccination. While reported data show an estimated 32.3% of eligible attendees received at least one dose of the HPV vaccine from the pilot start in 2016 to the end of 2024 [8], this is an underestimate [14] and other data sources suggest uptake has been higher, with 65% of GBMSM self-reporting vaccination in an online community survey from 2022 [15]. A very small proportion of GBMSM (1.1% in 2024) have been reported to decline an offer of vaccination [8].

We report findings from multi-site surveillance of type-specific anal HPV infections among GBMSM attending SSHSs, to monitor programme impact on infection rates, and estimate vaccine effectiveness, as an early indicator of whether the programme will meet its aims.

## METHODS

### Study design and participants

Residual rectal swab samples from chlamydia and/or gonorrhoea testing were collected from individuals self-reporting as GBMSM aged up to 45 years attending specialist SHSs in England between 2017-2019 (Phase 1, i.e. at the start of the pilot HPV vaccination programme) and 2021-2024 (Phase 2, i.e. 3-6 years after the rollout of the national programme). The British Association for Sexual Health and HIV (BASHH) guidance recommends that sexually active GBMSM undergo three-site sampling (first-void urine, pharyngeal swab and rectal swab) for chlamydia and/or gonorrhoea screening [16]. Some SHSs obtain a pooled specimen that combines all three sites. A study conducted at a London SHS found high concordance between residual rectal swab specimens and pooled specimens (obtained for chlamydia and/or gonorrhoea testing) for HPV DNA detection [17]. Both rectal swabs and pooled specimens were therefore deemed acceptable sample types for this surveillance.

Sociodemographic variables (age, ethnicity, and Index of Multiple Deprivation (IMD)), indicators of sexual risk (HIV, AGW and other STI diagnoses, and HIV pre-exposure prophylaxis (PrEP) use), and HPV vaccinations as recorded in the GUMCAD STI Surveillance System [18] were extracted and linked to the residual specimen. GUMCAD is a national surveillance system with mandatory reporting of all STI tests, diagnoses and services from all 241 publicly funded SHSs in England. GUMCAD collects pseudonymised, patient-level data, including a unique patient identifier linking attendances by the same patient within a SHS (or umbrella SHS, i.e. a group of associated services) from 1st January 2008. Therefore, patients cannot be tracked between clinics. In England, IMD measures relative levels of deprivation by Lower-layer Super Output Areas (LSOA), >30,000 small areas or neighbourhoods in the country [19].

This surveillance was conducted as public health monitoring of the HPV vaccination programme: individual patient consent was not required. UKHSA has permission to handle these data for this purpose under Regulation 3 of The Health Service (Control of Patient Information) Regulations 2020 and Section 251 of the National Health Service Act 2006.

### Specimen and data collection

Five laboratories provided specimens from six clinics during Phase 1, and three laboratories provided specimens from four clinics in Phase 2 (Table 1). Laboratories were asked to send a minimum volume of 1mL of the residual specimen (after the STI test had been completed) and for the sample to be kept refrigerated but not frozen. Sites were asked to send specimens unbiased by test results (i.e. both positive and negative samples). Patient identifiable information was removed from the sample tubes which were labelled with only a unique study identifier and barcode (provided by UKHSA, in duplicate pairs). Specimens were sent to the Virus Reference Department (VRD) laboratory at UKHSA Colindale.

**Table 1.** Sociodemographic characteristics, recorded vaccination status, and other stratifications of interest for GBMSM included in Phase 1 and Phase 2 surveys.

|  | **Phase 1 (2017-2019)**<br>**(n=3,291)** | **Phase 2 (2021-2024)**<br>**(n=2,496)** |
| --- | --- | --- |
| **Number of samples by laboratory and clinic** |  |  |
| Brighton & Sussex University Hospitals NHS Trust | 603 (18.3%) | 0 (0%) |
| Imperial College Healthcare NHS Trust (London) | 566 (17.2%) | 0 (0%) |
| Manchester Royal Infirmary | 766 (23.3%) | 0 (0%) |
| The Doctors Laboratory (London) | 715 (21.7%) | 655 (26.2%) |
| Newcastle UKHSA Regional Laboratory (Freeman Hospital) | 641 (19.5%) | 654 (26.2%) |
| Bristol UKHSA South West Regional Laboratory | 0 (0%) | 515 (20.6%) |
| 56 Dean St (London) | 0 (0%) | 672 (26.9%) |
| **Age, years** |  |  |
| ≤25 | 840 (25.5%) | 597 (23.9%) |
| 26-30 | 783 (23.8%) | 615 (24.6%) |
| 31-35 | 695 (21.1%) | 598 (24.0%) |
| 36-40 | 566 (17.2%) | 441 (17.7%) |
| 41-45 | 407 (12.4%) | 245 (9.8%) |
| **Ethnicity** |  |  |
| Asian | 145 (4.4%) | 163 (6.5%) |
| Black | 116 (3.5%) | 108 (4.3%) |
| Mixed | 162 (4.9%) | 166 (6.7%) |
| Other ethnic groups | 175 (5.3%) | 159 (6.4%) |
| White | 2358 (71.6%) | 1749 (70.1%) |
| Not specified | 335 (10.2%) | 151 (6.0%) |
| **IMD quintile<sup>1</sup>** |  |  |
| Q1 (most deprived) | 736 (22.4%) | 467 (18.7%) |
| Q2 | 1051 (31.9%) | 845 (33.9%) |
| Q3 | 736 (22.4%) | 582 (23.3%) |
| Q4 | 525 (16.0%) | 346 (13.9%) |
| Q5 (least deprived) | 243 (7.4%) | 256 (10.3%) |
| **Vaccination status** |  |  |
| Unvaccinated (including vaccinated on/after attendance) | 1821 (55.3%) | 1300 (52.1%) |
| Vaccinated (partial)<sup>2</sup> | 516 (15.7%) | 256 (10.3%) |
| Vaccinated (full)<sup>3</sup> | 954 (29.0%) | 940 (37.7%) |
| **Time since vaccination** |  |  |
| Within 1 year since initiation | 954 (29.0%) | 326 (13.1%) |
| Within 2 years since initiation | 479 (14.6%) | 160 (6.4%) |
| Within 3 years since initiation | 11 (0.3%) | 146 (5.8%) |
| >3 years since initiation | 0 (0%) | 516 (20.7%) |
| **History of bacterial STI** | 2149 (65.3%) | 1767 (70.8%) |
| **HIV status: Person living with HIV** | 681 (20.7%) | 189 (7.6%) |
| **History of genital warts diagnosis (first episode)** | 340 (10.3%) | 161 (6.5%) |
| **PrEP use at attendance<sup>4</sup>** | 255 (7.7%) | 589 (23.6%) |

<sup>1</sup> Index of Multiple Deprivation based on lower-layer super output (LSOA) areas in England
<sup>2</sup> 1 dose (at least 30 days prior to attendance)
<sup>3</sup> 2 or more doses (at least 30 days prior to attendance)
<sup>4</sup> Starting or continuing a daily or event based PrEP regimen and/or prescription

Laboratories completed a form for each specimen and affixed (or scanned) the second barcode/study identifier label to this form, providing the patient identifier, clinic code, attendance date in clinic, the date the sample was taken and patient age (in years) at that date. These forms were sent to the study team at UKHSA Colindale where these data were used for linkage to GUMCAD. After extraction of GUMCAD variables, all forms from the clinic and the clinic code and patient identifiers were deleted before the HPV test result was added to the study records using the study identifier. GBMSM were defined as unvaccinated if they had no HPV vaccine doses recorded in GUMCAD prior to the attendance date or if they had received a dose on or after the attendance date or less than 30 days prior to the attendance date, and as vaccinated if they had one or more doses recorded as given at least 30 days prior to the attendance date.

Residual rectal swabs were tested for type-specific HPV DNA to detect 13 high-risk types (HPV16, 18, 31, 33, 35, 39, 45, 51, 52, 56, 58, 59 and 68), six possible high-risk types (HPV26, 53, 66, 70, 73 and 82) and two low-risk types (HPV6 and 11) as per the International Agency for Research on Cancer (IARC) classifications [20], using an in-house multiplex PCR and Luminex based genotyping test [21]. PCR results were reported as inadequate if the samples were negative for both HPV DNA and the housekeeping gene, pyruvate dehydrogenase.

### Statistical analysis

The target sample size was 3,700 in each survey period, to detect a statistically significant difference in the prevalence of vaccine-type HPV infection among unvaccinated vs vaccinated GBMSM and allowing for up to 20% attrition (loss of samples due to not being retrieved at local laboratories and for inadequate test results).

Descriptive analysis was conducted for the prevalence of individual vaccine types and HPV type groupings, as follows: HPV16/18; HPV6/11; HPV6/11/16/18 (i.e. quadrivalent vaccine types); HPV31/33/45; other high-risk HPV types (HPV35/39/51/52/56/58/59/68); a grouping of non-vaccine-preventable/affected HPV (NVP-HPV) (HPV26/53/66/70/73), a group of low-risk HPV types not affected by HPV vaccination [22]. Results were stratified by age group, ethnicity, IMD quintile and indicators of sexual risk for both vaccination survey periods.

The change in prevalence of HPV (individual types and groupings) in Phase 1 and Phase 2 was analysed using a log-binomial regression model to calculate crude prevalence ratios (PRs). The 95% confidence intervals corresponding to the PRs were calculated using the Wald test. Adjusted PRs (aPRs) were also calculated adjusted for age, living with HIV, and bacterial STI diagnoses. Similarly, vaccine effectiveness was assessed by calculating PRs and aPRs by recorded vaccination status (i.e. vaccinated vs unvaccinated GBMSM). Analyses were conducted in Stata (version 18.0).

## RESULTS

### Patient sociodemographics and sample characteristics

A total of 7,368 specimens were received from laboratories across both survey periods (3,891 during the Phase 1 survey and 3,477 during the Phase 2 survey), and 7,073 (96%) were matched to GUMCAD data. 1,004 (14%) were excluded as ineligible (non-GBMSM or outside of age parameters). A total of 6,069 specimens (3,492 Phase 1 specimens and 2,577 Phase 2 specimens) were eligible for HPV testing. 282 specimens had an inadequate result.

Results were analysed for 5,787 specimens collected from eligible GBMSM (3,291 Phase 1 and 2,496 Phase 2). The characteristics of GBMSM included in the Phase 1 and Phase 2 surveys are summarised in Table 1. Previous STI diagnoses were common with >65% having had a bacterial STI diagnosis, and 7-10% a AGW diagnosis. The proportion of GBMSM living with HIV was higher within Phase 1 (20.7% vs 7.6%), while PrEP use was higher in Phase 2 (23.6% vs 7.7%). The proportions with HPV vaccination recorded was similar in both periods, although the average time since vaccination was longer in Phase 2.

### Changes in the prevalence of vaccine-type HPV infections

The prevalence of HPV16/18 infection was 36% (95% CI 28-44) lower in Phase 2 (11.8% (95% CI 10.6-13.2)) compared to Phase 1 (19.5% (95% CI 18.1-20.9)) (Table 2). HPV6/11/16/18 prevalence was 23% (95% CI 17-29) lower in Phase 2 (26.9% (95% CI 25.2-28.7) than in Phase 1 (36.0% (95% CI 34.4-37.7)). The prevalence of all individual and grouped vaccine-types, except for HPV11, was lower in Phase 2 compared to Phase 1 (Figure 1).

**Figure 1.**
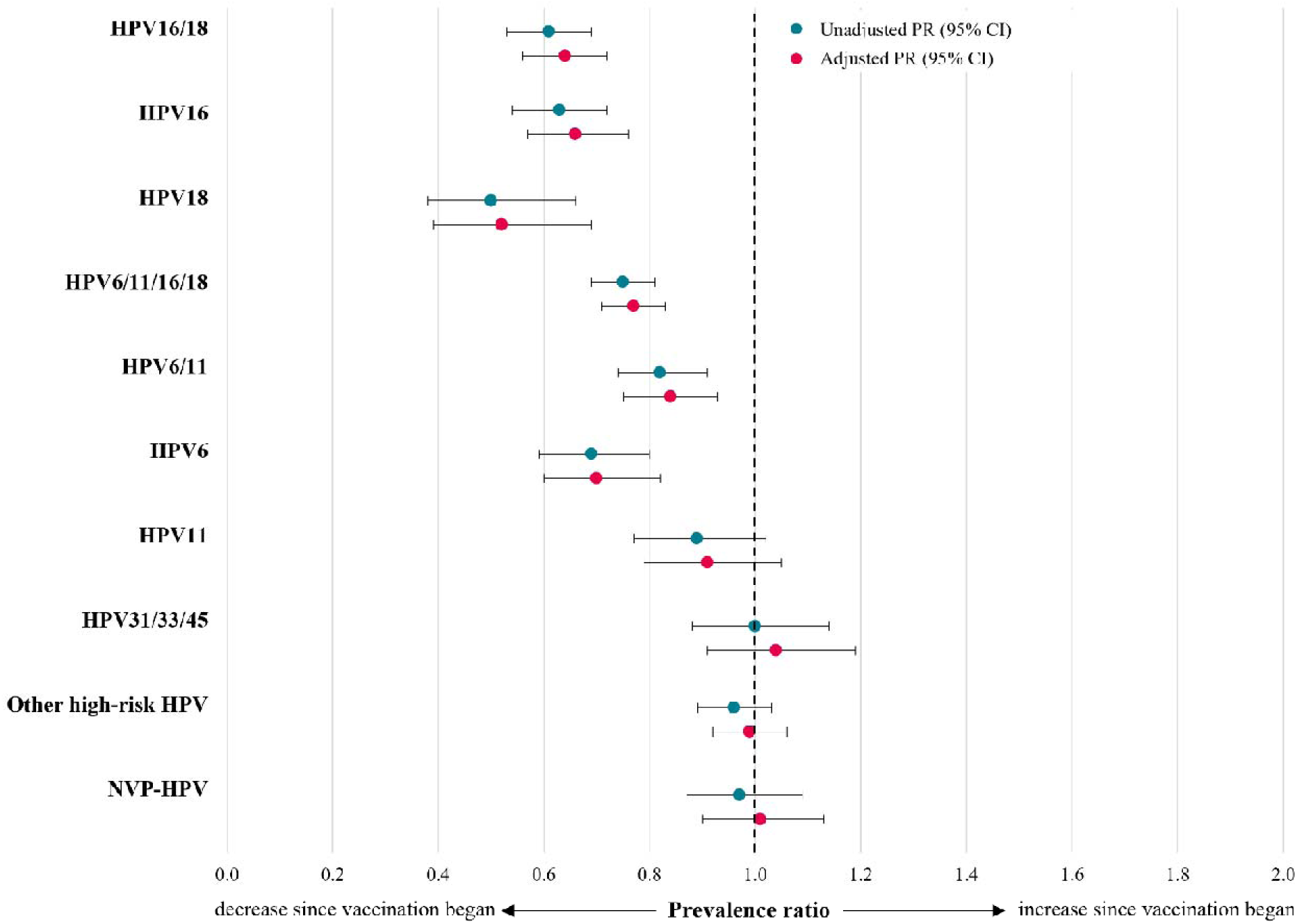
Unadjusted and adjusted PRs comparing individual and grouped anal vaccine-type HPV infection among GBMSM in Phase 1 versus Phase 2 survey periods.

**Table 2.** Comparison of type-specific HPV prevalence between the Phase 1 and Phase 2 survey periods.

|  | *Phase 1 (n=3,291)* | *Phase 2 (n=2,496)* |  |  |  |  |
| --- | --- | --- | --- | --- | --- | --- |
|  | **Prevalence (95% CI)** | **Prevalence (95% CI)** | **PR (95% CI)** | **p-value** | **aPR<sup>1</sup> (95% CI)** | **p-value** |
| **HPV16** | 15.5 (14.3-16.8) | 9.7 (8.6-10.9) | 0.63 (0.54-0.72) | <0.001 | 0.66 (0.57-0.76) | <0.001 |
| **HPV18** | 5.3 (4.6-6.2) | 2.7 (2.1-3.4) | 0.50 (0.38-0.66) | <0.001 | 0.52 (0.39-0.69) | <0.001 |
| **HPV16/18** | 19.5 (18.1-20.9) | 11.8 (10.6-13.2) | 0.61 (0.53-0.69) | <0.001 | 0.64 (0.56-0.72) | <0.001 |
| **HPV6** | 12.7 (11.6-13.9) | 8.7 (7.6-9.9) | 0.69 (0.59-0.80) | <0.001 | 0.70 (0.60-0.82) | <0.001 |
| **HPV11** | 13.3 (12.2-14.5) | 11.9 (10.6-13.2) | 0.89 (0.77-1.02) | 0.095 | 0.91 (0.79-1.05) | 0.19 |
| **HPV6/11** | 21.7 (20.3-23.1) | 17.7 (16.3-19.3) | 0.82 (0.74-0.91) | <0.001 | 0.84 (0.75-0.93) | 0.001 |
| **HPV6/11/16/18** | 36.0 (34.4-37.7) | 26.9 (25.2-28.7) | 0.75 (0.69-0.81) | <0.001 | 0.77 (0.71-0.83) | <0.001 |
| **HPV31** | 3.0 (2.5-3.7) | 1.8 (1.3-2.4) | 0.60 (0.42-0.85) | 0.0040 | 0.65 (0.45-0.92) | 0.017 |
| **HPV33** | 3.3 (2.8-4.0) | 4.4 (3.7-5.3) | 1.33 (1.03-1.72) | 0.030 | 1.45 (1.11-1.89) | 0.0070 |
| **HPV45** | 8.0 (7.1-9.0) | 7.9 (6.9-9.1) | 0.99 (0.83-1.18) | 0.90 | 1.01 (0.84-1.21) | 0.93 |
| **HPV31/33/45** | 13.5 (12.3-14.7) | 13.5 (12.2-14.9) | 1.00 (0.88-1.14) | 0.96 | 1.04 (0.91-1.19) | 0.58 |
| **Other (non-16/18/31/33/45) high-risk HPV** | 36.4 (34.8-38.1) | 34.9 (33.0-36.8) | 0.96 (0.89-1.03) | 0.23 | 0.99 (0.92-1.06) | 0.69 |
| **NVP-HPV** | 18.7 (17.4-20.1) | 18.2 (16.7-19.8) | 0.97 (0.87-1.09) | 0.64 | 1.01 (0.90-1.13) | 0.89 |

<sup>1</sup> Adjusted for age, history of bacterial STI and HIV status

The prevalence of HPV31/33/45 (types closely related to HPV16/18) did not differ between survey periods: 13.5% (95% CI 12.3-14.7) in Phase 1 and 13.5% (95% CI 12.2-14.9) in Phase 2. Similarly, there was no difference in the prevalence of the other high-risk HPV group or the NVP-HPV group (Table 2).

Differences in HPV16/18 and HPV6/11/16/18 prevalence between Phase 1 and Phase 2 were greater in the youngest age groups. Among ≤25-year-old GBMSM, HPV16/18 prevalence was 66% (95% CI 52-76) lower and HPV6/11/16/18 prevalence was 42% (95% CI 30-51) lower in Phase 2 compared to Phase 1 (Table 3). Changes in the prevalence of vaccine-types decreased with increasing age. There was no difference in vaccine-type HPV prevalence between Phase 1 and Phase 2 among 41–45-year-old GBMSM.

**Table 3.** Comparison of vaccine-type HPV prevalence between the Phase 1 and Phase 2 survey periods, by age group.

|  | *Phase 1* | *Phase 2* |  |  |  |  |
| --- | --- | --- | --- | --- | --- | --- |
|  | **Prevalence (95% CI)** | **Prevalence (95% CI)** | **PR (95% CI)** | **p-value** | **aPR<sup>1</sup> (95% CI)** | **p-value** |
| **Age group** |  |  |  |  |  |  |
| **≤25** | *n=840* | *n=597* |  |  |  |  |
| HPV16/18 | 16.9 (14.4-19.6) | 5.7 (4.0-7.9) | 0.34 (0.24-0.48) | <0.001 | 0.34 (0.24-0.48) | <0.001 |
| HPV6/11/16/18 | 34.8 (31.5-38.1) | 20.4 (17.3-23.9) | 0.59 (0.49-0.71) | <0.001 | 0.58 (0.49-0.70) | <0.001 |
| NVP-HPV | 13.3 (11.1-15.8) | 13.7 (11.1-16.8) | 1.03 (0.79-1.34) | 0.83 | 1.00 (0.77-1.30) | 0.99 |
| **26-30** | *n=783* | *n=615* |  |  |  |  |
| HPV16/18 | 20.4 (17.7-23.4) | 12.4 (9.9-15.2) | 0.60 (0.47-0.78) | <0.001 | 0.65 (0.50-0.84) | <0.001 |
| HPV6/11/16/18 | 37.0 (33.6-40.5) | 29.3 (25.7-33.0) | 0.79 (0.68-0.92) | 0.0030 | 0.82 (0.70-0.95) | 0.010 |
| NVP-HPV | 17.2 (14.7-20.1) | 17.4 (14.5-20.6) | 1.01 (0.80-1.27) | 0.94 | 1.01 (0.80-1.28) | 0.92 |
| **31-35** | *n=695* | *n=598* |  |  |  |  |
| HPV16/18 | 20.3 (17.4-23.5) | 13.0 (10.4-16.0) | 0.64 (0.50-0.83) | <0.001 | 0.65 (0.50-0.84) | <0.001 |
| HPV6/11/16/18 | 35.4 (31.8-39.1) | 27.1 (23.6-30.8) | 0.77 (0.65-0.90) | 0.0020 | 0.77 (0.65-0.92) | 0.0030 |
| NVP-HPV | 20.7 (17.8-23.9) | 18.7 (15.7-22.1) | 0.90 (0.72-1.13) | 0.37 | 0.96 (0.76-1.20) | 0.71 |
| **36-40** | *n=566* | *n=441* |  |  |  |  |
| HPV16/18 | 21.6 (18.2-25.2) | 14.7 (11.6-18.4) | 0.68 (0.52-0.90) | 0.0070 | 0.74 (0.56-0.99) | 0.043 |
| HPV6/11/16/18 | 35.9 (31.9-40.0) | 29.7 (25.5-34.2) | 0.83 (0.69-0.99) | 0.041 | 0.88 (0.73-1.06) | 0.18 |
| NVP-HPV | 23.3 (19.9-27.0) | 22.0 (18.2-26.2) | 0.94 (0.75-1.19) | 0.62 | 0.95 (0.75-1.21) | 0.68 |
| **41-45** | *n=407* | *n=245* |  |  |  |  |
| HPV16/18 | 18.7 (15.0-22.8) | 17.1 (12.6-22.5) | 0.92 (0.65-1.29) | 0.62 | 0.98 (0.69-1.40) | 0.93 |
| HPV6/11/16/18 | 38.1 (33.3-43.0) | 31.4 (25.7-37.6) | 0.83 (0.66-1.03) | 0.091 | 0.87 (0.69-1.09) | 0.23 |
| NVP-HPV | 22.9 (18.9-27.2) | 23.3 (18.1-29.1) | 1.02 (0.76-1.36) | 0.90 | 1.13 (0.84-1.51) | 0.43 |

<sup>1</sup> Adjusted for age, history of bacterial STI and HIV status

### Vaccine effectiveness

The prevalence of vaccine-type HPV infections was similar amongst GBMSM recorded as vaccinated (1 or more doses) and GBMSM with no recorded vaccination, overall (Table 4) and within each survey (Supplementary Material).

**Table 4.** Comparison of type-specific HPV prevalence in vaccinated versus unvaccinated GBMSM.

|  | *Unvaccinated*<br>*GBMSM (n=3,121)* | *Vaccinated<sup>1</sup>*<br>*GBMSM (n=2,666)* |  |  |  |  |
| --- | --- | --- | --- | --- | --- | --- |
|  | **Prevalence (95%**<br>**CI)** | **Prevalence (95%**<br>**CI)** | **PR (95% CI)** | **p-value** | **aPR<sup>2</sup> (95% CI)** | **p-value** |
| **HPV16** | 12.4 (11.3-13.6) | 13.7 (12.4-15.0) | 1.10 (0.96-1.25) | 0.17 | 0.97 (0.85-1.11) | 0.67 |
| **HPV18** | 4.3 (3.6-5.0) | 4.1 (3.4-5.0) | 0.97 (0.76-1.24) | 0.80 | 0.86 (0.67-1.11) | 0.26 |
| **HPV16/18** | 15.8 (14.5-17.1) | 16.6 (15.2-18.1) | 1.05 (0.94-1.18) | 0.40 | 0.94 (0.83-1.06) | 0.32 |
| **HPV6** | 10.5 (9.5-11.6) | 11.5 (10.3-12.7) | 1.09 (0.94-1.27) | 0.24 | 1.02 (0.87-1.18) | 0.84 |
| **HPV11** | 12.2 (11.0-13.4) | 13.3 (12.0-14.7) | 1.09 (0.96-1.25) | 0.19 | 0.98 (0.85-1.13) | 0.77 |
| **HPV6/11** | 19.3 (17.9-20.7) | 20.8 (19.3-22.4) | 1.08 (0.97-1.19) | 0.16 | 0.98 (0.88-1.09) | 0.66 |
| **HPV6/11/16/18** | 31.1 (29.5-32.7) | 33.3 (31.5-35.1) | 1.07 (0.99-1.16) | 0.070 | 0.98 (0.91-1.06) | 0.59 |
| **HPV31** | 2.3 (1.8-2.9) | 2.7 (2.2-3.4) | 1.20 (0.87-1.66) | 0.26 | 1.01 (0.72-1.41) | 0.96 |
| **HPV33** | 3.2 (2.6-3.9) | 4.5 (3.7-5.4) | 1.39 (1.07-1.80) | 0.013 | 1.13 (0.86-1.47) | 0.38 |
| **HPV45** | 7.4 (6.5-8.4) | 8.7 (7.6-9.8) | 1.17 (0.98-1.39) | 0.077 | 1.03 (0.86-1.23) | 0.75 |
| **HPV31/33/45** | 12.2 (11.1-13.4) | 15.0 (13.7-16.4) | 1.23 (1.08-1.40) | 0.0020 | 1.05 (0.92-1.20) | 0.45 |
| **Other (non-16/18/31/33/45) high-risk HPV** | 32.2 (30.6-33.9) | 39.9 (38.1-41.8) | 1.24 (1.16-1.33) | <0.001 | 1.10 (1.02-1.18) | 0.0090 |
| **NVP-HPV** | 16.1 (14.8-17.4) | 21.4 (19.8-23.0) | 1.33 (1.20-1.48) | <0.001 | 1.13 (1.02-1.27) | 0.025 |

<sup>1</sup> 1 or more doses (at least 30 days prior to attendance)
<sup>2</sup> Adjusted for age, history of bacterial STI and HIV status

The prevalence of HPV31/33/45 (types closely related to HPV16/18) was higher in GBMSM recorded as vaccinated (15.0%; 95% CI 13.7-16.4) than in those with no recorded vaccination (12.2%; 95% CI 11.1-13.4) (PR 1.23; 95% CI 1.08-1.40). However, there was no difference after adjustment for potential confounders, i.e. age, HIV status, and history of bacterial STI (aPR 1.05; 95% CI 0.92-1.20). There was a higher prevalence of other high-risk HPV in vaccinated GBMSM (39.9%; 95% CI 38.1-41.8) compared to those with no recorded vaccination (32.2%; 95% CI 30.6-33.9) (aPR 1.10; 95% CI 1.02-1.18). The prevalence of NVP-HPV infection was also substantially higher in vaccinated GBMSM (21.4%; 95% CI 19.8-23.0 compared to unvaccinated GBMSM (16.1%; 95% CI 14.8-17.4) (PR 1.33; 95% CI 1.20-1.48 and aPR 1.13; 95% CI 1.02-1.27).

There was a difference in HPV6/11/16/18 infection between recorded vaccinated versus unvaccinated ≤25-year-old GBMSM, (aPR 0.82; 95% CI 0.69-0.97) (Table 5).

**Table 5.** Comparison of vaccine-type HPV prevalence in vaccinated versus unvaccinated GBMSM, by age group.

|  | *Unvaccinated GBMSM* | *Vaccinated<sup>1</sup> GBMSM* |  |  |  |  |
| --- | --- | --- | --- | --- | --- | --- |
|  | **Prevalence (95% CI)** | **Prevalence (95% CI)** | **PR (95% CI)** | **p-value** | **aPR<sup>2</sup> (95% CI)** | **p-value** |
| **Age group** |  |  |  |  |  |  |
| **≤25** | *n=878* | *n=559* |  |  |  |  |
| HPV16/18 | 12.5 (10.4-14.9) | 11.8 (9.3-14.8) | 0.94 (0.71-1.25) | 0.68 | 0.85 (0.64-1.14) | 0.29 |
| HPV6/11/16/18 | 29.8 (26.8-33.0) | 27.2 (23.5-31.1) | 0.91 (0.77-1.08) | 0.28 | 0.82 (0.69-0.97) | 0.024 |
| NVP-HPV | 11.8 (9.8-14.2) | 16.1 (13.1-19.4) | 1.36 (1.05-1.77) | 0.021 | 1.21 (0.92-1.58) | 0.17 |
| **26-30** | *n=777* | *n=621* |  |  |  |  |
| HPV16/18 | 17.5 (14.9-20.4) | 16.1 (13.3-19.2) | 0.92 (0.73-1.16) | 0.49 | 0.83 (0.65-1.06) | 0.14 |
| HPV6/11/16/18 | 32.3 (29.0-35.7) | 35.3 (31.5-39.2) | 1.09 (0.94-1.27) | 0.24 | 0.99 (0.85-1.15) | 0.87 |
| NVP-HPV | 14.9 (12.5-17.6) | 20.3 (17.2-23.7) | 1.36 (1.08-1.71) | 0.0090 | 1.23 (0.97-1.57) | 0.082 |
| **31-35** | *n=674* | *n=619* |  |  |  |  |
| HPV16/18 | 16.8 (14.0-19.8) | 17.1 (14.2-20.3) | 1.02 (0.80-1.30) | 0.86 | 0.96 (0.75-1.23) | 0.74 |
| HPV6/11/16/18 | 30.3 (26.8-33.9) | 33.0 (29.3-36.8) | 1.09 (0.93-1.28) | 0.30 | 1.02 (0.87-1.20) | 0.80 |
| NVP-HPV | 19.0 (16.1-22.2) | 20.7 (17.6-24.1) | 1.09 (0.87-1.36) | 0.45 | 0.98 (0.78-1.22) | 0.84 |
| **36-40** | *n=485* | *n=522* |  |  |  |  |
| HPV16/18 | 16.5 (13.3-20.1) | 20.5 (17.1-24.2) | 1.24 (0.96-1.61) | 0.10 | 1.16 (0.89-1.52) | 0.28 |
| HPV6/11/16/18 | 32.2 (28.0-36.5) | 34.1 (30.0-38.3) | 1.06 (0.89-1.26) | 0.52 | 0.99 (0.83-1.18) | 0.92 |
| NVP-HPV | 17.9 (14.6-21.6) | 27.2 (23.4-31.2) | 1.52 (1.20-1.92) | <0.001 | 1.29 (1.02-1.64) | 0.033 |
| **41-45** | *n=307* | *n=345* |  |  |  |  |
| HPV16/18 | 17.6 (13.5-22.3) | 18.6 (14.6-23.1) | 1.05 (0.76-1.46) | 0.75 | 0.90 (0.64-1.25) | 0.52 |
| HPV6/11/16/18 | 31.6 (26.4-37.1) | 39.1 (33.9-44.5) | 1.24 (1.00-1.53) | 0.047 | 1.12 (0.90-1.38) | 0.31 |
| NVP-HPV | 21.5 (17.0-26.5) | 24.3 (19.9-29.2) | 1.13 (0.85-1.50) | 0.39 | 0.97 (0.73-1.29) | 0.86 |

<sup>1</sup> 1 or more doses (at least 30 days prior to attendance)
<sup>2</sup> Adjusted for age, history of bacterial STI and HIV status

Results from sensitivity analysis (Supplementary Material) restricting to the consistent sites only (Newcastle and The Doctors Laboratory) did not differ substantially from the main analysis.

## DISCUSSION

These surveillance data provide the first multi-site estimates of anal HPV infection in GBMSM attending SHSs in England and are the first evidence of the impact of HPV vaccination in this population. Eight years after the introduction of targeted HPV vaccination for GBMSM, the prevalence of high-risk types HPV16/18 in GBMSM has declined by 36% and the prevalence of HPV6/11/16/18 has declined by 23%. Results by age group indicate that these decreases were confined to the three youngest age groups (≤25-, 26-30- and 31-35-year-old GBMSM). The observed reductions in HPV infection prevalence between the Phase 1 and Phase 2 survey periods predict declines in HPV-related disease among GBMSM, including AGW and anal cancers in the future. There was no evidence of any differences in the prevalence of closely related types HPV31/33/45 and other (non-16/18/31/33/45) high-risk HPV types between survey periods.

There was a lack of association between HPV infection prevalence and recorded vaccination status, with the exception of ≤25-year-old GBMSM, among whom the prevalence of HPV6/11/16/18 was 18% lower in vaccinated versus unvaccinated individuals. Measurement error in the form of underreporting of vaccinations has been estimated to be as high as 30% in some London services [14], which will have resulted in underestimation of vaccine effectiveness. Being recorded as vaccinated was associated with a higher risk of HPV, as evidenced by the 30% higher prevalence of NVP-HPV (a molecular indicator of sexual behaviour risk for HPV transmission) in GBMSM recorded as vaccinated. If GBMSM receiving vaccination were truly those at higher risk and therefore more likely to be infected with HPV, this would have biased vaccine effectiveness estimates toward the null while strengthening impact measures. A third factor that has likely reduced apparent vaccine effectiveness is herd protection, from both the adolescent and targeted GBMSM programmes. Herd immunity from the GBMSM programme, which may be estimated at up to 20% as per previous modelling work [10], would also impact estimates of vaccine effectiveness via decreased HPV prevalence in the unvaccinated subgroup. The changes in prevalence of vaccine types amongst GBMSM attending SHSs may also be partly due to some herd protection from the national adolescent (female only for this time period) vaccination programme, which has reduced the prevalence of HPV16/18 in young women markedly since 2008 [9]. Under 25-year-old GBMSM are most likely to benefit from herd protection from the female programme due to increased sexual mixing at younger ages [10], particularly those attending during the Phase 2 survey period. Finally, this lack of effect may be age-related, as older GBMSM are more likely to have had multiple sexual partners and prior exposure to multiple HPV types (including the vaccine types), resulting in reduced or absent vaccine effectiveness in this subgroup. Incomplete vaccination recording at sexual health services and selection bias in vaccination delivery could similarly bias any estimates of vaccine effectiveness using this method for the mpox and 4CMenB vaccination programmes (also delivered in SHSs), and this will need to be taken into account when evaluating these new vaccination programmes.

A smaller reduction in prevalence was to be expected in this population compared to adolescents, due to the higher risk of prior infection with vaccine-type infections. Also, given the higher prevalence in GBMSM and the high sensitivity of HPV DNA testing, even for low viral loads [23], some infections detected may have been transient HPV infections or viral depositions in the anal canal that vaccination will prevent from infecting the epithelial cells. In clinical trials, vaccine efficacy against the detection of any vaccine-type HPV DNA was 27% in GBMSM, considerably lower than against persistent infection or disease [24].

Our findings are based on ecological observations. - There were changes in sexual health service use over our study period, and there may have been changes in sexual behaviour that could have affected HPV prevalence independently to vaccination. We found differences in HIV status (i.e. the proportion of GBMSM living with HIV) between the two survey periods, but no difference in the prevalence of NVP-HPV or bacterial STI diagnoses. Over this period, there was a scale-up in PrEP use among GBMSM and evidence of increased incidence of bacterial STIs among PrEP users [25]. These changes would likely have driven increases in high-risk HPV prevalence of both vaccine and non-vaccine HPV types. We adjusted for variables that could be related to risk for HPV (age, history of bacterial STI and HIV status) in log-binomial regression models to explore the impact of these possible confounding effects. There were several changes in the provision of, or attendance at, SHSs for GBMSM, including a shift to online postal self-sampling services and evidence of unmet needs for in-person SHS access, which would have likely increased high-risk HPV prevalence in face-to-face attendees [26, 27]. Additionally, there were changes in the sites participating in Phase 1 and Phase 2. However, the vaccine-type infection prevalence in the Phase 1 survey was similar across sites. Our surveillance started concurrently with the pilot programme and so does not provide a true baseline for HPV prevalence pre-vaccination. A 2010-2012 study conducted in a London clinic, which used the same assay and informed the national programme, estimated HPV16/18 prevalence at 18.4% and HPV6/11/16/18 prevalence at 32.5% [11], only slightly lower than our corresponding Phase 1 estimates (and within the 95% CIs). Finally, there may have been other, unmeasured confounders that affected the estimated association between vaccination and changes in HPV prevalence between the two periods.

The UK is the only country with a national vaccination programme for GBMSM, but several other countries have issued recommendations to address the same health inequality and have reported on vaccination impact in this population and our findings are largely aligned with similar real-world studies. In Scotland, where GBMSM vaccination started at the same time as in England, vaccine-type HPV prevalence decreased from 44.6% pre-GBMSM-vaccination (2016-2017) to 34.7% in the latest (third) post-vaccination survey period (2021) [28]. In Canada, where some provinces offer vaccination for GBMSM under 26 years of age, 16-30 year old GBMSM had a 27% lower prevalence of HPV6/11/16/18 compared to unvaccinated GBMSM [29]. In the United States, where HPV vaccination is recommended for GBMSM aged up to 26 years, an analysis of HPV vaccine effectiveness in GBMSM aged 18-45 years old attending three sexual health clinics found high (87%) vaccine effectiveness against HPV6/11/16/18 prevalence among GBMSM aged 18–26 years old who were vaccinated at age <18 [30]. Lower (approximately 30%) vaccine effectiveness was estimated among GBMSM aged 27–45 years old who were vaccinated aged 18-26 or at least 2 years prior to specimen collection. Finally, in Australia, where some states have implemented catch-up vaccination for GBMSM aged up to and including 26 years, a study conducted at one sexual health clinic found reductions of 69% in HPV16/18 prevalence and 76% in HPV6/11/16/18 prevalence 4-5 years after the start of vaccination in young 16-20 year old GBMSM [31].

The incremental benefits of the targeted HPV vaccination programme for GBMSM at the population level will gradually diminish as cohorts of vaccinated adolescents age into adulthood and increasing proportions of GBMSM will have been eligible for vaccination in the routine school programme. The long natural history of progression from HPV infection to anal cancer means that vaccination has a limited impact on disease in the short term, as pre-existing infections will present for several decades to come. In the interim, older GBMSM and those living with HIV who remain at highest risk may still benefit from secondary prevention measures such as anal screening. The consideration of an anal cancer screening programme for GBMSM should be informed by these novel data on the epidemiology of HPV infection in this population.

## CONCLUSION

Our analysis provides the first epidemiological evidence of the impact of targeted HPV vaccination for GBMSM in England. The findings suggest that the aim of reducing the risk of HPV-related disease, and reducing HPV-related health inequality, in this population is being achieved. The vaccine effectiveness findings may be due to the combined effects of measurement error (underreporting of vaccination in STI surveillance data), herd protection, and bias towards vaccination of higher risk GBMSM. The evidence supports continuing the programme in England until the national adolescent HPV vaccination programme has been established long enough to offer this group direct protection, and potentially beyond that to provide catch-up vaccination for those who missed their vaccination in school and remain unprotected.

## Data Availability

All data produced in the present study are available upon reasonable request to the authors

## Acknowledgments

The authors are grateful to the participating laboratories who provided residual specimens for testing. The authors also thank the GUMCAD STI Surveillance System team for providing expertise on the GUMCAD dataset. They are also grateful to Anja Anderson and Farida Abdulkadir for assisting with data collection and processing. Finally, the authors thank Tom Pink from UKHSA Knowledge and Library Services for his help with the Research in Context literature search.

## Contributors

This surveillance was initiated and designed by KS and MC. MC was responsible for the sample collection and data management. HM is the guarantor for the GUMCAD STI Surveillance System. SB and KP performed the HPV testing. MC conducted the statistical analysis, data interpretation and wrote a first draft of the manuscript. KS, MH and CEF provided advice on and contributed to the study design, statistical analysis and interpretation of results. All authors revised the manuscript and approved the final version of this manuscript.

