## Supplementary Material for "Impact of the HPV vaccination programme on anal HPV infection in gay, bisexual and other men who have sex with men: multi-site enhanced surveillance study in specialist sexual health services in England"

Supplementary Table 1 Prevalence of vaccine-type HPV infection in Phase 1 and Phase 2 survey periods, by vaccination status

|  | ***Phase 1 (n=3,291)*** | | | | | |
| --- | --- | --- | --- | --- | --- | --- |
|  | ***Unvaccinated GBMSM (n=1,821)*** | ***Vaccinated******^[1]^ GBMSM (n=1,470)*** |  |  |  |  |
|  | **Prevalence (95% CI)** | **Prevalence (95% CI)** | **PR (95% CI)** | **p-value** | **Adjusted****^[2]^ PR (95% CI)** | **p-value** |
| **HPV16/18** | 19.1 (17.3-21.0) | 19.9 (17.9-22.1) | 1.04 (0.91-1.2) | 0.55 | 0.94 (0.82-1.09) | 0.41 |
| **HPV6/11/16/18** | 35.3 (33.1-37.6) | 36.9 (34.5-39.5) | 1.05 (0.95-1.15) | 0.33 | 0.95 (0.86-1.04) | 0.27 |
|  | ***Phase 2 (n=2,496)*** | | | | | |
|  | ***Unvaccinated GBMSM (n=1,300)*** | ***Vaccinated^1^ GBMSM (n=1,196)*** |  |  |  |  |
|  | **Prevalence (95% CI)** | **Prevalence (95% CI)** | **PR (95% CI)** | **p-value** | **Adjusted^2^ PR (95% CI)** | **p-value** |
| **HPV16/18** | 11.2 (9.5-13.0) | 12.5 (10.7-14.6) | 1.12 (0.91-1.39) | 0.28 | 0.97 (0.78-1.21) | 0.80 |
| **HPV6/11/16/18** | 25.2 (22.8-27.6) | 28.8 (26.3-31.5) | 1.15 (1.01-1.31) | 0.038 | 1.06 (0.93-1.21) | 0.41 |

Supplementary Table 2 Sociodemographic characteristics, recorded vaccination status, and other stratifications of interest for GBMSM included in Phase 1 and Phase 2 surveys: The Doctors’ Laboratory and Newcastle sites only

|  | **Phase 1 (2017-2019)** | **Phase 2 (2021-2024)** |
| --- | --- | --- |
|  | ***(n=1,356)*** | ***(n=1,309)*** |
| **Number of samples by laboratory and clinic** |  |  |
| The Doctors Laboratory (London) | 715 (52.7%) | 655 (50.0%) |
| Newcastle UKHSA Regional Laboratory (Freeman Hospital) | 641 (47.3%) | 654 (50.0%) |
| **Age, years** |  |  |
| ≤25 | 402 (29.6%) | 343 (26.2%) |
| 26-30 | 326 (24.0%) | 347 (26.5%) |
| 31-35 | 276 (20.4%) | 296 (22.6%) |
| 36-40 | 227 (16.7%) | 214 (16.3%) |
| 41-45 | 125 (9.2%) | 109 (8.3%) |
| **Ethnicity** |  |  |
| Asian | 70 (5.2%) | 93 (7.1%) |
| Black | 40 (2.9%) | 61 (4.7%) |
| Mixed | 60 (4.4%) | 84 (6.4%) |
| Other ethnic groups | 82 (6.0%) | 79 (6.0%) |
| White | 898 (66.2%) | 893 (68.2%) |
| Not specified | 206 (15.2%) | 99 (7.6%) |
| **IMD quintile^[3]^** |  |  |
| Q1 (most deprived) | 261 (19.2%) | 258 (19.7%) |
| Q2 | 407 (30.0%) | 425 (32.5%) |
| Q3 | 363 (26.8%) | 338 (25.8%) |
| Q4 | 200 (14.7%) | 171 (13.1%) |
| Q5 (least deprived) | 125 (9.2%) | 117 (8.9%) |
| **Vaccination status** |  |  |
| Unvaccinated (including vaccinated on/after attendance) | 705 (52.0%) | 697 (53.2%) |
| Vaccinated (partial)^[4]^ | 180 (13.3%) | 138 (10.5%) |
| Vaccinated (full)^[5]^ | 471 (34.7%) | 474 (36.2%) |
| **Time since vaccination** |  |  |
| Within 1 year since initiation | 356 (26.3%) | 170 (13.0%) |
| Within 2 years since initiation | 274 (20.2%) | 68 (5.2%) |
| Within 3 years since initiation | 11 (0.8%) | 60 (4.6%) |
| >3 years since initiation | 0 (0.0%) | 294 (22.5%) |
| **History of bacterial STI** | 863 (63.6%) | 890 (68.0%) |
| **History of HIV diagnosis (HIV status)** | 229 (16.9%) | 101 (7.7%) |
| **History of genital warts diagnosis (first episode)** | 128 (9.4%) | 70 (5.3%) |
| **PrEP use at attendance^[6]^** | 96 (7.1%) | 415 (31.7%) |

Supplementary Table 3 Comparison of type-specific HPV prevalence between the Phase 1 and Phase 2 survey periods: The Doctors’ Laboratory and Newcastle sites only

|  | ***Phase 1 (n=1,356)*** | ***Phase 2 (n=1,309)*** |  |  |  |  |
| --- | --- | --- | --- | --- | --- | --- |
|  | **Prevalence (95% CI)** | **Prevalence (95% CI)** | **PR (95% CI)** | **p-value** | **Adjusted^[7]^ PR (95% CI)** | **p-value** |
| **HPV16** | 14.8 (13.0-16.8) | 8.7 (7.2-10.4) | 0.59 (0.47-0.73) | <0.001 | 0.61 (0.49-0.76) | <0.001 |
| **HPV18** | 5.1 (4.0-6.4) | 2.6 (1.8-3.6) | 0.51 (0.34-0.76) | 0.0010 | 0.51 (0.34-0.77) | 0.0010 |
| **HPV16/18** | 18.6 (16.5-20.8) | 10.8 (9.1-12.6) | 0.58 (0.48-0.70) | <0.001 | 0.59 (0.49-0.72) | <0.001 |
| **HPV6** | 12.0 (10.3-13.9) | 9.1 (7.6-10.8) | 0.76 (0.60-0.95) | 0.014 | 0.76 (0.6-0.95) | 0.017 |
| **HPV11** | 15.1 (13.3-17.1) | 11.0 (9.4-12.8) | 0.73 (0.60-0.89) | 0.0020 | 0.73 (0.6-0.89) | 0.0020 |
| **HPV6/11** | 21.7 (19.5-24.0) | 18.0 (16.0-20.2) | 0.83 (0.71-0.97) | 0.019 | 0.83 (0.71-0.97) | 0.016 |
| **HPV16/18/6/11** | 36.0 (33.4-38.6) | 26.8 (24.4-29.3) | 0.75 (0.66-0.84) | <0.001 | 0.75 (0.66-0.84) | <0.001 |
| **HPV31** | 2.9 (2.1-3.9) | 1.6 (1.0-2.4) | 0.56 (0.33-0.94) | 0.029 | 0.61 (0.36-1.04) | 0.068 |
| **HPV33** | 3.2 (2.3-4.2) | 3.6 (2.6-4.7) | 1.13 (0.75-1.70) | 0.55 | 1.18 (0.78-1.78) | 0.44 |
| **HPV45** | 6.9 (5.6-8.4) | 8.3 (6.8-9.9) | 1.19 (0.91-1.55) | 0.20 | 1.21 (0.92-1.59) | 0.17 |
| **HPV31/33/45** | 12.5 (10.8-14.3) | 12.9 (11.1-14.8) | 1.04 (0.85-1.26) | 0.73 | 1.06 (0.87-1.3) | 0.56 |
| **Other (non-16/18/31/33/45) high-risk HPV** | 35.0 (32.4-37.6) | 34.5 (31.9-37.1) | 0.99 (0.89-1.09) | 0.79 | 0.99 (0.89-1.09) | 0.79 |
| **NVP-HPV** | 17.1 (15.1-19.2) | 18.4 (16.3-20.6) | 1.08 (0.91-1.27) | 0.38 | 1.07 (0.91-1.26) | 0.43 |

Supplementary Table 4 Comparison of type-specific HPV prevalence in vaccinated versus unvaccinated GBMSM: The Doctors’ Laboratory and Newcastle sites only

|  | ***Unvaccinated GBMSM (n=1,402)*** | ***Vaccinated^[8]^ GBMSM (n=1,263)*** |  |  |  |  |
| --- | --- | --- | --- | --- | --- | --- |
|  | **Prevalence (95% CI)** | **Prevalence (95% CI)** | **PR (95% CI)** | **p-value** | **Adjusted^[9]^ PR (95% CI)** | **p-value** |
| **HPV16** | 10.8 (9.2-12.5) | 13.0 (11.2-15.0) | 1.21 (0.98-1.48) | 0.077 | 1.05 (0.85-1.30) | 0.65 |
| **HPV18** | 3.4 (2.5-4.4) | 4.4 (3.4-5.7) | 1.32 (0.90-1.93) | 0.15 | 1.16 (0.78-1.72) | 0.45 |
| **HPV16/18** | 13.6 (11.9-15.5) | 16.0 (14.0-18.1) | 1.17 (0.98-1.41) | 0.085 | 1.04 (0.86-1.25) | 0.70 |
| **HPV6** | 10.1 (8.5-11.8) | 11.2 (9.5-13) | 1.11 (0.89-1.38) | 0.35 | 1.09 (0.86-1.37) | 0.48 |
| **HPV11** | 13.6 (11.8-15.5) | 12.6 (10.8-14.5) | 0.93 (0.76-1.13) | 0.46 | 0.84 (0.69-1.03) | 0.096 |
| **HPV6/11** | 19.8 (17.8-22) | 20.0 (17.8-22.3) | 1.01 (0.86-1.17) | 0.94 | 0.93 (0.79-1.09) | 0.36 |
| **HPV16/18/6/11** | 30.1 (27.7-32.6) | 33.0 (30.4-35.7) | 1.10 (0.98-1.23) | 0.11 | 1.01 (0.90-1.13) | 0.92 |
| **HPV31** | 2.1 (1.4-3.0) | 2.5 (1.7-3.5) | 1.19 (0.72-1.96) | 0.50 | 1.02 (0.60-1.71) | 0.95 |
| **HPV33** | 2.7 (1.9-3.7) | 4.1 (3.1-5.4) | 1.52 (1.01-2.29) | 0.046 | 1.11 (0.73-1.68) | 0.63 |
| **HPV45** | 7.1 (5.8-8.5) | 8.2 (6.7-9.8) | 1.15 (0.89-1.51) | 0.29 | 1.01 (0.77-1.33) | 0.94 |
| **HPV31/33/45** | 11.3 (9.7-13.0) | 14.3 (12.4-16.3) | 1.26 (1.04-1.54) | 0.021 | 1.07 (0.87-1.31) | 0.52 |
| **Other (non-16/18/31/33/45) high-risk HPV** | 30.9 (28.5-33.4) | 39.0 (36.3-41.7) | 1.26 (1.14-1.40) | <0.001 | 1.13 (1.01-1.26) | 0.026 |
| **NVP-HPV** | 14.9 (13.1-16.9) | 20.9 (18.7-23.3) | 1.40 (1.19-1.65) | <0.001 | 1.20 (1.01-1.42) | 0.035 |

1. 1 or more doses (at least 30 days prior to attendance)
2. Adjusted for age, history of bacterial STI and HIV status
3. Index of Multiple Deprivation based on lower-layer super output (LSOA) areas in England
4. 1 dose (at least 30 days prior to attendance)
5. 2 or more doses (at least 30 days prior to attendance)
6. Starting or continuing a daily or event based PrEP regimen and/or prescription
7. Adjusted for age, history of bacterial STI and HIV status
8. 1 or more doses (at least 30 days prior to attendance)
9. Adjusted for age, history of bacterial STI and HIV status
